# Transcobalamin Receptor Autoantibodies in Central Vitamin B12 Deficiency

**DOI:** 10.1101/2023.08.21.23294253

**Authors:** John V. Pluvinage, Thomas Ngo, Christopher M. Bartley, Aaron Bodansky, Bonny D. Alvarenga, Kelsey C. Zorn, Camille Fouassier, Colin Zamecnik, Adrian McCann, Trung Huynh, Weston Browne, Asritha Tubati, Sravani Kondapavulur, Mark S. Anderson, Ari J. Green, Ralph Green, Vanja Douglas, Martineau Louine, Bruce Cree, Stephen Hauser, William Seeley, Brandon B. Holmes, James A. Wells, Serena Spudich, Shelli Farhadian, Prashanth Ramachandran, Leslie Gillum, Chadwick M. Hales, Bryan Smith, Avindra Nath, Gina Suh, Eoin P. Flanagan, Jeffrey M. Gelfand, Joseph L. DeRisi, Samuel J. Pleasure, Michael R. Wilson

## Abstract

Vitamin B12 is critical for hematopoiesis and myelination.^1^ Deficiency can cause neurologic deficits including loss of coordination, spasticity, and cognitive decline.^2,3,4^ However, diagnosis relies on vitamin B12 measurement in the blood which may not accurately reflect levels in the brain. Here, we discovered an autoimmune cause of vitamin B12 deficiency restricted to the central nervous system (CNS), termed autoimmune B12 central deficiency (ABCD). Using programmable phage display, we identified an autoantibody targeting the transcobalamin receptor (CD320) in a patient with progressive tremor, ataxia, and scanning speech. Patient immunoglobulins impaired cellular uptake of vitamin B12 *in vitro*. Despite normal serum levels, vitamin B12 was nearly undetectable in her cerebrospinal fluid (CSF). Immunosuppressive treatment and high-dose systemic vitamin B12 supplementation were associated with increased CSF B12 levels and clinical improvement. Autoantibodies targeting the same epitope of CD320 were identified in 7 other patients with neurologic deficits of unknown etiology and in 6 percent of healthy controls. In 132 paired serum and CSF samples, detection of anti-CD320 in the blood predicted B12 deficiency in the brain. These findings elucidate a new autoimmune cause of metabolic neurologic disease that may be amenable to immunomodulatory treatment and/or nutritional supplementation.

## Case Report

A woman in her 60s presented with 5 weeks of progressive bilateral lower extremity pain, difficulty speaking, ataxia, and tremor (Case 1; Fig. 1A). Her neurologic exam was notable for scanning speech, a rubral tremor of the head and hands, diminished vibratory sensation in her feet, dysmetria, and truncal ataxia. MRI of the brain revealed bilateral symmetric T2-weighted hyperintensities in the dorsal brainstem with gadolinium enhancement of the superior cerebellar peduncles (Fig. 1B). There were no lesions on MRI of the cervical spine. Lumbar puncture yielded non-inflammatory CSF (2 white blood cells, 2 red blood cells, total protein of 54 mg/dL, glucose of 91 mg/dL, zero oligoclonal bands, and a normal IgG index of 0.6; Fig. 1C; Table S1). Clinical autoimmune encephalopathy antibody testing was negative, including testing for AQP4 antibodies. MOG antibody testing 4 years later was also negative. A broad rheumatologic workup demonstrated elevated titers of antinuclear, antiphospholipid, and anti-double stranded DNA antibodies. A repeat MRI 11 days later demonstrated progression of disease, and she was empirically treated with pulse dose glucocorticoids.

**Figure 1.**
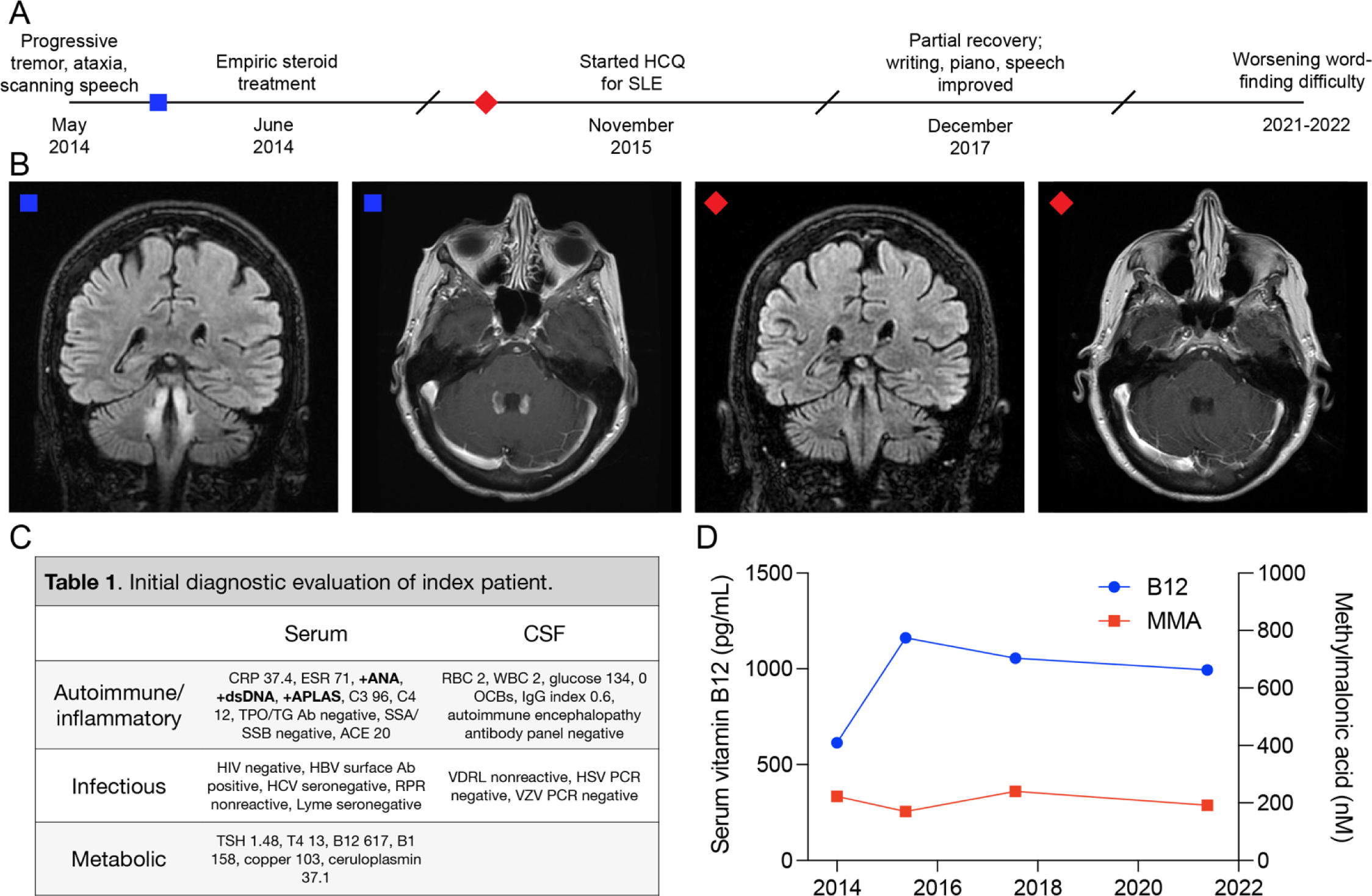
Clinical course and initial diagnostic investigation of Case 1. Panel A shows the clinical timeline from initial presentation to stabilization. The blue square corresponds to the timepoint of pre-treatment brain MRI sequences. The red diamond corresponds to the timepoint of post-treatment brain MRI sequences. Panel B shows coronal slices of T2-weighted fluid-attenuated inversion recovery (FLAIR) MRI and axial slices of T1-weighted post-gadolinium MRI before (left two images) and 9 months after (right two images) steroid treatment. T2-weighted hyperintensities in the superior cerebellar peduncles, midbrain tegmentum, dentate nuclei, chief sensory nuclei of cranial nerve V, and rubrospinal tracts persist, but contrast enhancement decreases after immunosuppression. Panel C shows the initial diagnostic tests sent from serum and CSF. Bolded items are abnormal. Panel D shows normal serum vitamin B12 and methylmalonic acid levels throughout the clinical course.

Her neurologic exam incrementally improved over the subsequent 3 months, and a repeat brain MRI showed an interval decrease in gadolinium enhancement. Eighteen months after her initial presentation, she was formally diagnosed with systemic lupus erythematosus (SLE) and treated with hydroxychloroquine (HCQ). Her functional status slowly recovered over the next 3 years: her speech improved, her handwriting became legible, and she returned to playing the piano. Surveillance brain MRI showed resolution of contrast-enhancement but persistent T2 hyperintensities. Although her initial neurologic deficits stabilized, she experienced gradually worsening word-finding difficulty. Formal neuropsychologic assessment demonstrated mild impairments in processing speed, fluency, and naming. Blood tests for reversible causes of dementia were normal, including serum vitamin B12 levels between 614 and 1162 pg/mL and methylmalonic acid (MMA, a metabolite that accumulates downstream of B12 deficiency) levels between 170 and 240 nM (Fig. 1D).

## Methods

The patient was enrolled in a research study to detect novel autoantibodies in suspected neuroinflammatory disease (IRB 13-12236). Programmable phage immunoprecipitation sequencing (PhIP-seq)^5^ was used to screen for autoantibodies against a library containing ∼730,000 peptide sequences tiling approximately 50,000 human proteoforms (https://github.com/derisilab-ucsf/PhIP-PND-2018).^6^ CSF was incubated with the phage display library, and antibody-bound phage were immunoprecipitated, sequenced, and analyzed using a previously described pipeline. Candidate autoantigens were validated using human embryonic kidney 293T (HEK293T) cell-based overexpression assays, CRISPR-Cas9-mediated knockout in primary human brain endothelial cells, and split-luciferase binding assays.

An *in vitro* holotranscobalamin trafficking assay was adapted to evaluate whether CSF autoantibodies interfered with cellular uptake of B12.^7^ Total vitamin B12 and holotranscobalamin were measured using commercially available enzyme-linked immunosorbent assays. MMA was measured by gas chromatography mass spectrometry. Autoantibodies were measured in the serum of a healthy control cohort (IRB 22-36117) and in both the serum and CSF of a cohort of patients with multiple sclerosis or other neurologic diseases (IRB 14-15278).

## Results

We screened for candidate autoantigens using PhIP-Seq. Among enriched antigens meeting a false discovery rate cutoff of <5%, CD320 was the top cell surface hit, a characteristic consistent with a pathogenic autoantibody (Table S2).^8^ Antibodies in the patient’s CSF selectively immunoprecipitated a peptide in the extracellular domain of CD320, distant from the low-density lipoprotein receptor type A (LDLR-A) motifs that form the transcobalamin binding interface (Fig. 2A).^9–10^ Sequential alanine mutagenesis of the peptide further narrowed the critical region for autoantibody binding to a 15 amino acid epitope (Fig. S1A). Overexpression of CD320 in a human HEK293T cell-based assay increased CSF antibody binding (Fig. 2B). Immunoreactivity was also observed in the serum (Fig. S1B). Conversely, CRISPR-Cas9-mediated knockout of CD320 in primary human brain endothelial cells ablated CSF antibody binding at the live cell surface (Fig. 2C). These findings confirm that CD320 is the cell-surface target of an autoantibody present in the serum and CSF.

**Figure 2.**
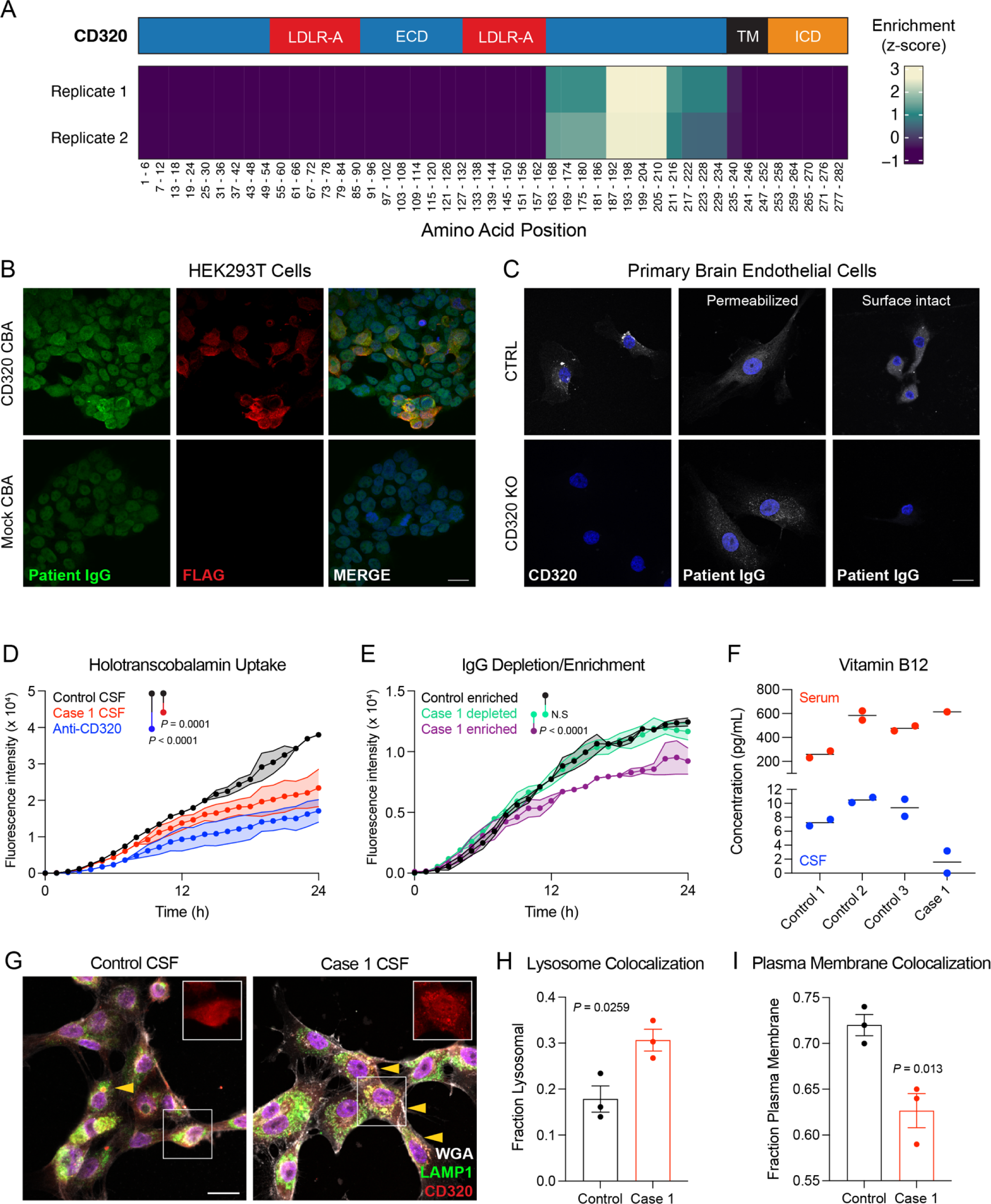
Discovery and validation of a functional transcobalamin receptor autoantibody. Panel A shows an epitope map of CD320 peptides aligned to the full-length gene (NP_057663.1; coverage is divided into five amino acid bins). CSF antibodies in both technical replicates enrich a common epitope in the extracellular domain of CD320 centered at amino acids 187-210. The blue region represents the extracellular domain (ECD), the red regions represent low-density lipoprotein receptor type A (LDLR-A) domains within the ECD, the black region represents the transmembrane (TM) domain, and the orange region represents the intracellular domain (ICD). Panel B shows increased immunoreactivity of patient CSF at 1:25 dilution in HEK293T cells overexpressing a FLAG-tagged CD320 construct. Nuclear immunoreactivity was expected given high titer anti-nuclear antibodies in the index patient. Scale bar, 20 microns. Panel C shows control and CD320-knockout primary human brain endothelial cells stained with a commercial CD320 antibody (left column) or patient CSF at 1:25 dilution (middle and right columns). CSF immunoreactivity persists in permeabilized CD320 KO cells but is absent in surface-intact CD320 KO cells. These results suggest that multiple autoantibodies exist in the index patient’s CSF, but CD320 is the only cell surface autoantigen. Scale bar, 20 microns. Panel D shows impaired holotranscobalamin uptake in cells treated with Case 1 CSF (red) or a commercial CD320 antibody (blue) compared to cells treated with healthy control CSF (black) (N=3, paired one-way ANOVA with Tukey’s multiplicity correction, mean +/- standard error). Depletion of immunoglobulins from Case 1’s CSF eliminated its inhibitory effect on holotranscobalamin uptake, as shown in Panel E. In contrast, enriched CSF immunoglobulin displayed potent inhibition (N=3, paired one-way ANOVA with Tukey’s multiplicity correction, mean +/- standard error). Panel F shows vitamin B12 concentration in serum (red) and CSF (blue) of three healthy controls and Case 1. Absolute CSF vitamin B12 concentration and the ratio of CSF to serum vitamin B12 are low in Case 1, suggesting a CNS-restricted vitamin B12 deficiency. Panel G shows representative images of brain endothelial cells treated with control or patient CSF, followed by staining for CD320 (red), a plasma membrane marker (WGA; gray), and a lysosomal marker (LAMP1; green). Treatment with patient CSF increased CD320-lysosome colocalization (yellow arrows, scale bar = 20 microns) and decreased CD320-plasma membrane colocalization, as quantified in Panels H and I (N=3; two-sided t-test; mean +/- standard error).

CD320, also known as the transcobalamin receptor, is enriched in endothelial cells at the blood-brain barrier (Fig. S2A-C) and mediates the cellular uptake and transcytosis of transcobalamin-bound vitamin B12 (holotranscobalamin) into the CNS.^11–12^ Incubation of HEK293T cells with patient CSF, but not healthy control CSF devoid of anti-CD320, inhibited holotranscobalamin uptake comparable to treatment with a commercial anti-CD320 polyclonal antibody (Fig. 2D, S3A-D). Immunoglobulin depletion from patient CSF eliminated this inhibitory effect, and patient CSF had no effect on holotranscobalamin uptake in CD320-deficient cells (Fig. 2E, S3E-G). Immunostaining of human brain tissue demonstrated CSF antibody binding in CD31+ endothelial cells (Fig. S3H). Treatment of human brain endothelial cells with patient CSF increased lysosomal colocalization and decreased plasma membrane colocalization of CD320 (Fig. 2G-I). These findings demonstrate patient immunoglobulins inhibit the function of CD320 and suggest that anti-CD320 may impair vitamin B12 transport across the blood-brain barrier by depleting cell-surface receptor availability.

To interrogate a possible blood-brain barrier transport defect in this patient, we measured vitamin B12 in paired serum and CSF samples. In three healthy controls, the mean serum concentration of vitamin B12 was 439 pg/mL and the mean CSF concentration was 9.0 pg/mL, yielding a CSF to serum vitamin B12 ratio of 0.02, concordant with prior studies (Fig. 2F).^13,14^ In contrast, the patient’s serum vitamin B12 concentration was 617 pg/mL and CSF concentration was 1.6 pg/mL, yielding a CSF to serum vitamin B12 ratio of 0.003, nearly one order of magnitude lower than healthy controls. High-dose oral vitamin B12 supplementation was initiated, and the patient’s CSF B12 level increased to 4.8 pg/mL (Fig. S4E). After 9 months of treatment, she reported subjective improvements in mood and cognitive function.

We retrospectively analyzed PhIP-seq data from 254 individuals enrolled in a neuroinflammatory disease study. We identified 7 additional cases harboring a CSF anti-CD320 autoantibody targeting the same epitope as the antibody in the index patient (Cases 2-8; Fig. 3A; Fig. S4 A-C). These 7 individuals were clinically diverse, with a wide range of autoantibody titers (Fig. 3B, S4D). CSF from all seven cases functionally impaired holotranscobalamin uptake (Fig. 3C). Compared to antibody-negative healthy controls, B12 was low and MMA was elevated in the CSF of four cases (Fig. 3D). Holotranscobalamin, a sensitive serum marker of early subclinical vitamin B12 deficiency, was low in the CSF of all seven cases (Fig. 3E).^15^ In contrast, serum vitamin B12 was normal in all cases with paired blood samples (Table S3). Case 3 had T2-hyperintensities in the cerebellar peduncles and dorsal brainstem resembling the brain MRI of the index patient (Fig. 3F). Case 6 developed progressive ataxia, cognitive decline, and white matter disease in the setting of persistently elevated anti-CD320 titer on longitudinal CSF sampling (Fig. S4E). Case 8 presented with subacute combined degeneration of the dorsal columns and lateral corticospinal white matter tracts, a classic manifestation of systemic B12 deficiency (Fig. 3G). He partially recovered on B-cell depletion therapy and high-dose oral B12 supplementation.

**Figure 3.**
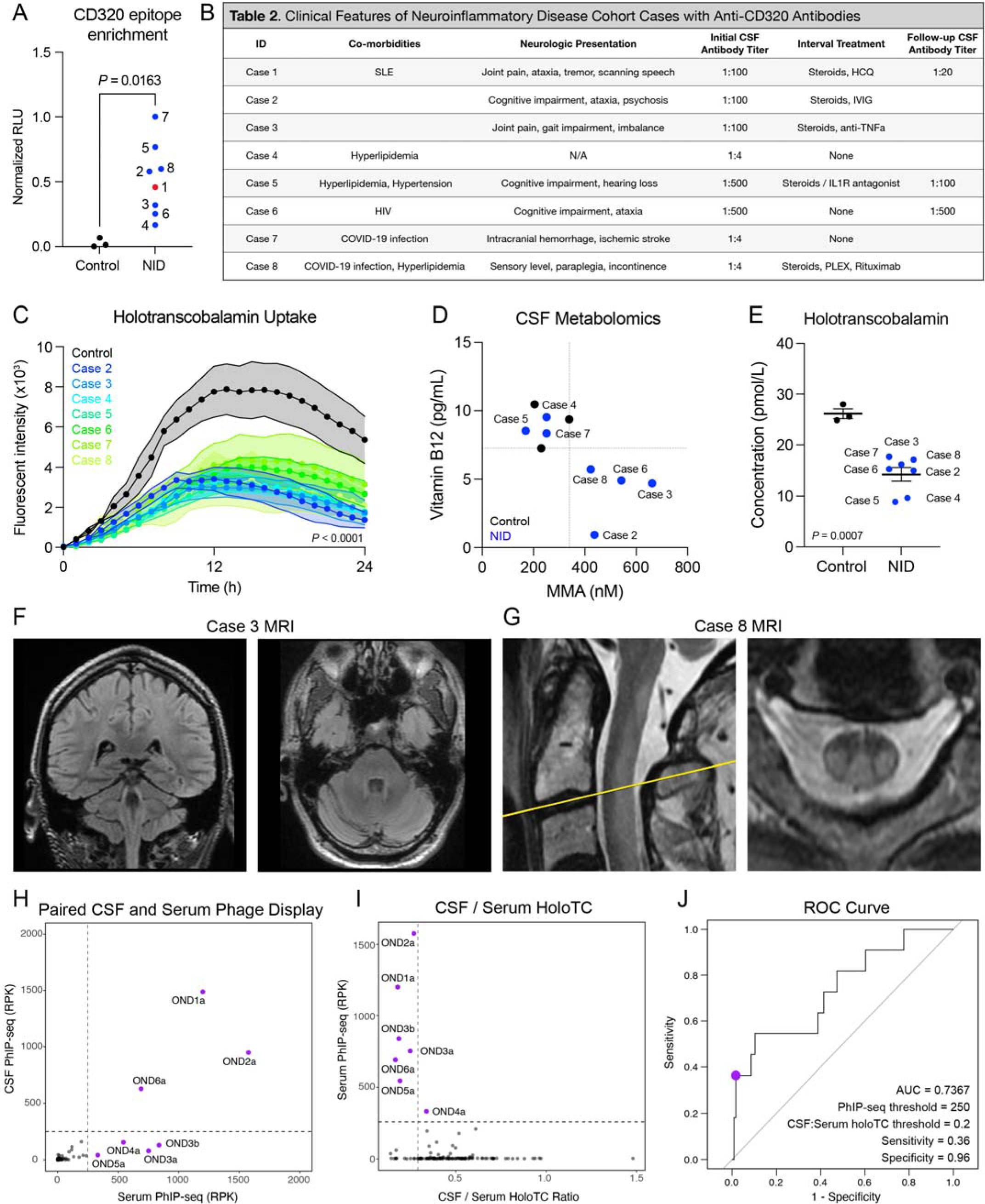
Retrospective identification of individuals with transcobalamin receptor autoantibodies. Panel A shows enrichment of a CD320 peptide by CSF immunoglobulin from 3 healthy controls (black) and 8 individuals enrolled in a neuroinflammatory disease study (NID, blue), including the index patient (red), using a split-luciferase binding assay (two-sided t-test). Panel B shows the clinical characteristics of individuals harboring CSF anti-CD320 autoantibodies. Autoantibody titer was determined by cell-based assay. Panel C shows holotranscobalamin uptake by cells treated with CSF from the 7 additional cases, each demonstrating functional inhibition compared to treatment with healthy control CSF (N=3, one-way ANOVA with Tukey’s multiplicity correction, mean +/- standard error). Panel D shows total vitamin B12 (y-axis) and MMA (x-axis) concentration in CSF from the 7 cases enrolled in a neuroinflammatory disease study (blue dots) and 3 healthy controls (black dots). Dotted lines show the lowest CSF vitamin B12 or MMA level among healthy controls. Four cases exhibit low total vitamin B12 and elevated MMA levels in their CSF, suggestive of central vitamin B12 deficiency with downstream metabolic consequences. Panel E shows holotranscobalamin concentration in CSF from healthy controls and cases in the neuroinflammatory disease cohort (two-sided t-test; mean +/- standard error). Panel F shows coronal and axial cuts of the T2-weighted FLAIR MRI of Case 3. Hyperintense bilateral symmetric T2-FLAIR signal involving the cerebellar peduncles resembles the index patient. Panel G shows sagittal and axial cuts of the T2-weighted C-spine MRI for Case 8. Hyperintense signal involving the dorsal columns and lateral corticospinal white matter tracts is classic for subacute combined degeneration. Panel H shows CD320 enrichment of paired serum and CSF samples from 132 patients with multiple sclerosis. Enrichment determined by PhIP-seq, where RPK represents depth-normalized sequencing reads. Labeled samples exhibit CD320 enrichment equal to or greater than 250 RPK (dotted line). Panel I shows CSF / serum holotranscobalamin ratio vs serum anti-CD320 enrichment in multiple sclerosis patients. The CSF / serum holotranscobalamin threshold (0.2) was chosen based on previously reported reference ranges in each biofluid.^14,25^ Panel J is the receiver operator characteristic (ROC) curve showing the performance of serum anti-CD320 seropositivity for predicting a low CSF / serum holotranscobalamin level. At an RPK threshold of 250, anti-CD320 was 36% sensitive and 96% specific for predicting central B12 deficiency. RPK threshold was chosen to stratify seropositive versus seronegative and should not be used as a proxy for autoantibody titer.

Due to the presence of transcobalamin receptor autoantibodies in the CSF of a healthy control (Case 4), we screened sera from a cohort of 84 additional healthy controls and identified anti-CD320 antibodies targeting the same epitope as the antibody in the index patient in 5 individuals, yielding a seropositivity rate of 6 percent (Fig. S5A). To determine the clinical relevance of this finding, we leveraged a large cohort of 132 paired serum and CSF samples from patients with multiple sclerosis (N=123) or other neurologic diseases (N=9). Anti-CD320 autoantibodies were detected in 5.7% of this cohort, concordant with the seropositivity rate in the healthy cohort (Fig. 3H). High-titer CSF samples impaired holotranscobalamin uptake (Fig. S5B). Detection of anti-CD320 in the serum was 36% sensitive and 96% specific for a low CSF to serum holotranscobalamin ratio (Fig. 3I-J), and holotranscobalamin accumulated in the serum of seropositive individuals (Fig. S5C). Furthermore, anti-CD320 seropositivity predicted elevated CSF MMA, a metabolic marker of vitamin B12 deficiency, with 78% positive predictive value (Fig. S5D-F).

## Discussion

We identified an autoantibody targeting the transcobalamin receptor in a patient with progressive tremor, ataxia, and scanning speech. Anti-CD320 impaired cellular uptake of holotranscobalamin and was associated with low levels of vitamin B12 in the CSF despite normal levels in the blood. We retrospectively found autoantibodies reactive to the same epitope of CD320 in seven individuals enrolled in a neuroinflammatory disease study and in 6 percent of a healthy control cohort. Detection of anti-CD320 in the blood predicted vitamin B12 deficiency in the CSF. These findings support a model in which anti-CD320 impairs transport of vitamin B12 across the blood-brain barrier, leading to ABCD with varied clinical manifestations (Fig. S6A).

The neurologic spectrum of traditional systemic vitamin B12 deficiency is wide, ranging from peripheral neuropathy to encephalomyelopathy. Prior to the onset of these symptoms, individuals progress from replete to deficient through a state of inadequacy known as subclinical vitamin B12 deficiency.^16^ We detected transcobalamin receptor autoantibodies in a wide range of cases, including healthy controls, patients with other neurologic conditions (MS), and a patient with classic signs of vitamin B12 deficiency despite a normal serum B12 level (Case 8). This heterogeneity may reflect differences in anti-CD320 antibody avidity, the degree and duration of CSF vitamin B12 deficiency, and/or host factors conferring differential susceptibility to disease (Fig. S6B). Although not directly addressed in the present study, the high specificity and positive predictive value of anti-CD320 in the blood for detecting early markers of B12 deficiency in the CSF suggest that seropositive healthy controls may be at risk for ABCD. In contrast to neuronal surface autoantibodies in autoimmune encephalitides (e.g. NMDA receptor antibody or LGI1 antibody) that are directly neuromodulatory,^17,18,19^ CD320 autoantibodies may exert a second order effect on brain function by first decreasing CSF levels of vitamin B12 that then leads to insidious neurologic sequelae. This study suggests that the measurement of vitamin B12 metabolites in CSF should be considered for anti-CD320 seropositive patients with unexplained neurologic deficits.

All cases shared autoantibodies targeting a common epitope in the extracellular domain of the transcobalamin receptor (Pro183-Thr192). An autoantibody roughly mapping to a similar region (Thr169-Tyr229) has also been described in patients with cutaneous arteritis.^20^ Future studies will be necessary to determine whether neurologic and dermatologic manifestations of anti-CD320 autoantibodies coexist.

Transcobalamin receptor autoantibodies were detected in both CSF and serum. Autoantibody enrichment was markedly higher in the serum, suggesting a peripheral source. Nevertheless, these individuals displayed none of the hematologic manifestations of vitamin B12 deficiency. Similarly, systemic vitamin B12 deficiency frequently presents as an isolated neuropsychiatric syndrome without anemia, CNS-restricted pathology is found in CD320-deficient mice, and humans with CD320 mutations are hematologically normal despite methylmalonic aciduria.^21,22,23^ These findings suggest the brain is selectively vulnerable to deficiency or that alternative vitamin B12 uptake pathways exist in peripheral tissues.

Several patients improved after immunosuppression (Cases 1, 2, 3, and 8), and two of these patients (Cases 1 and 8) received concurrent high-dose systemic B12 supplementation. It is unclear whether immunosuppression mediated improvement in these cases via dampening of alternative neuroinflammatory mechanisms or via mitigation of anti-CD320 autoantibody production. The increased B12 concentration in post-treatment CSF from Case 1 suggests that high serum B12 levels may suffice to overcome BBB transport defects caused by anti-CD320.

This study has several limitations. First, without prospective recruitment and empiric B12 targeted treatment of seropositive patients, it is not possible to determine whether anti-CD320 autoantibodies primarily drive disease pathogenesis, exacerbate underlying pathology (“second hit”), or passively spectate. Second, anti-CD320 sensitivity and specificity calculations were based on measurements in a cohort of patients with other neurologic diseases (primarily MS) which may not accurately reflect the clinical relevance of the autoantibody in healthy individuals. Future studies will be necessary to evaluate whether anti-CD320 autoantibodies modulate clinical outcomes in the general population and in other neurologic diseases like MS. Finally, this study should not be used as a comprehensive clinical characterization of ABCD given the heterogeneity of disease and atypical features (e.g. cerebellar peduncle involvement in Cases 1 and 3) that distinguish it from classic systemic vitamin B12 deficiency.

These findings parallel cerebral folate deficiency syndrome, in which autoantibodies targeting folate receptors at the choroid plexus are associated with low 5-methyltetrahydrofolate levels in the CSF despite normal levels in the blood.^24^ Although it is unclear whether transcobalamin receptor autoantibodies contribute to the primary pathophysiology of neurologic deterioration in these patients, correction of central vitamin B12 deficiency may provide clinical benefit with minimal risk. Future studies will be necessary to determine the therapeutic effect of high dose vitamin B12 supplementation and/or immunomodulatory therapy.

## Data Availability

All data produced in the present study are available upon reasonable request to the authors.

## Acknowledgments

We thank Ms. Maham Zia and Mr. Ali Khazaei for sample processing, Ms. PeiXi Chen and Dr. Michael Haney for equipment and reagents, the ORIGINS/EPIC research team for patient recruitment and sample collection, NIMH (R01MH122471), NINDS (U01NS120836; R35NS111644), Department of Defense (W81XWH-21-1-0979), National Multiple Sclerosis Society, Valhalla Foundation and the Westridge Foundation for funding, and the research subjects and their families for participation in this study.

---

**Supplementary Figure 1.**
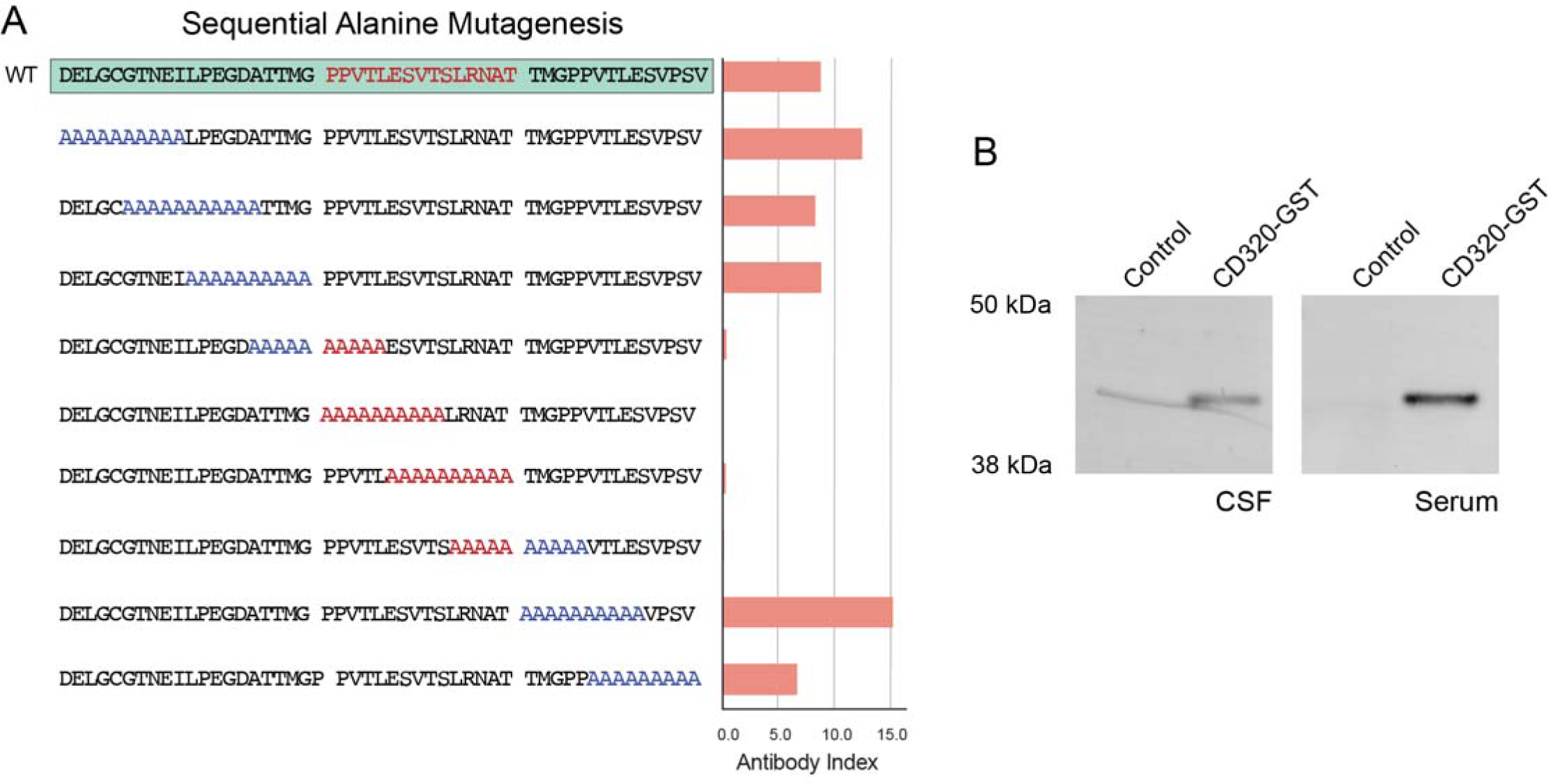
Mapping anti-CD320 epitope specificity. Panel A shows CSF immunoprecipitation of CD320 peptides mutagenized with alanine residues. Alanine substitution in a 15 amino acid stretch ablates peptide immunoprecipitation, clarifying this region as the target epitope for patient-derived anti-CD320 (Pro183-Thr192 of NP_057663.1). Panel B shows western blot confirmation of anti-CD320 antibodies in the index patient’s CSF and serum. Recombinant full-length CD320 tagged with glutathione s-transferase (GST) or a truncated version of CD320 lacking the auto-epitope were immunoblotted with patient CSF or serum. Both biofluids were selectively immunoreactive to CD320-GST containing the auto-epitope.

**Supplementary Figure 2.**
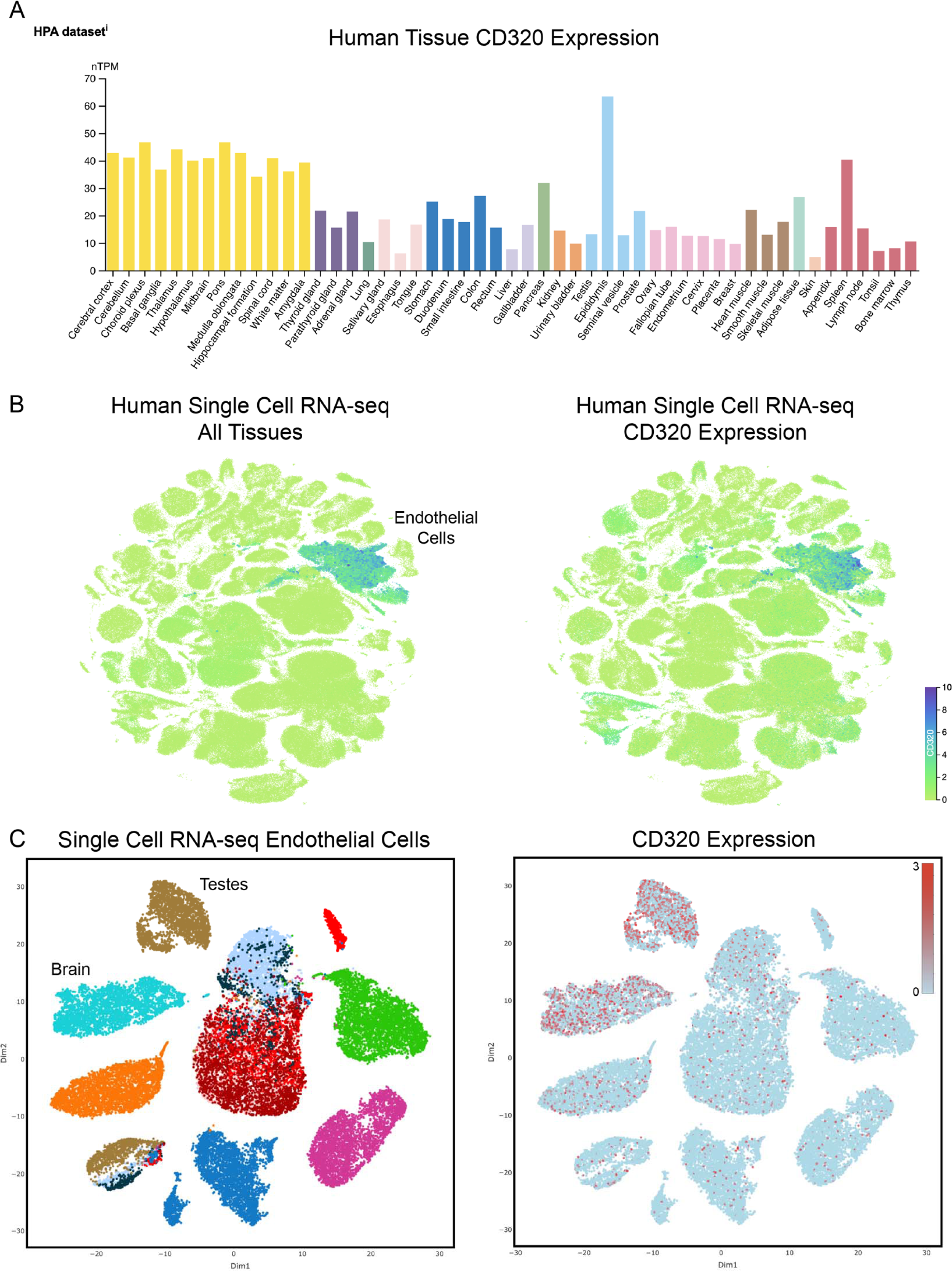
CD320 expression in human tissues and single cells. Panel A shows CD320 expression in multiple human tissues, as measured by bulk RNA-sequencing from the Human Protein Atlas. CD320 is broadly expressed across all tissues, and highly expressed in brain tissue. Panel B shows CD320 expression in multiple human tissues, as measured by single-cell RNA-sequencing from the Tabula Sapiens project. CD320 is highly expressed in endothelial cells. Panel C shows CD320 expression in endothelial cells from multiple human tissues, as measured by single-cell RNA-sequencing from the EC database. CD320 is expressed in all tissues but enriched in endothelial cells from brain and testes.

**Supplementary Figure 3.**
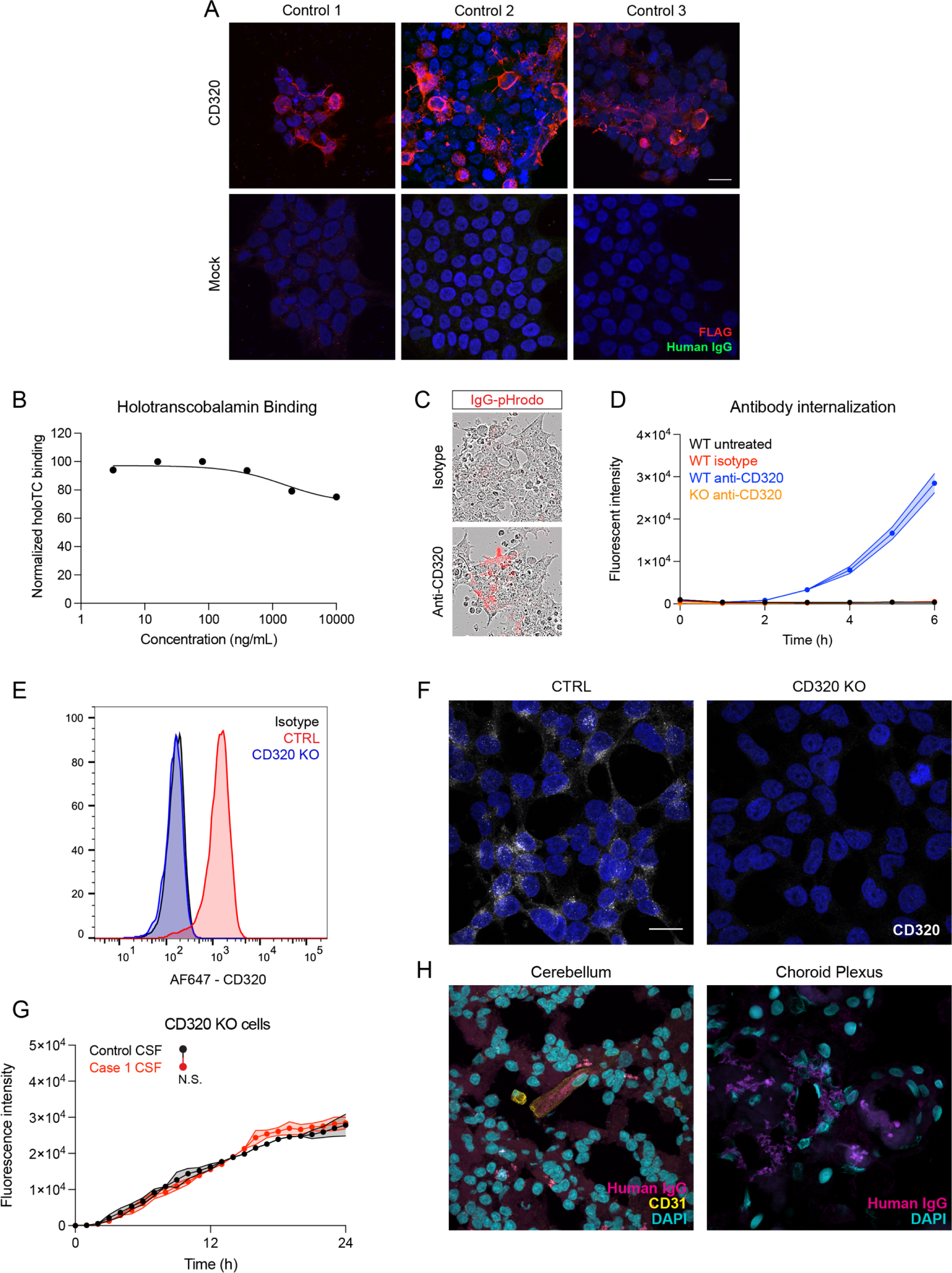
Immunoreactivity and mechanism of anti-CD320. Panel A shows immunoreactivity of healthy control CSF IgG for human cells overexpressing FLAG-tagged CD320. Intrathecal anti-CD320 autoantibodies were not detected in these healthy controls at a dilution of 1:4. Panel B demonstrates the lack of significant dose-dependent blockade of holotranscobalamin binding to the surface of HEK293T cells treated with a commercial anti-CD320 antibody. Panel C shows representative images of antibody internalization in cells treated with a commercial anti-CD320 antibody or an isotype control antibody. Panel D shows rapid internalization of a commercial anti-CD320 antibody by wild type HEK293T cells, but no internalization in isotype control treated cells or CD320 KO cells (N=3, mean +/- standard error). Panel E shows flow cytometric quantification of CD320 expression on control and CD320 KO HEK293T cells. Panel F shows immunocytochemical staining of control and CD320 KO HEK293T cells with a commercial CD320 antibody. Scale bar, 20 microns. Panel G shows no effect of Case 1 CSF on holotranscobalamin uptake compared to healthy control CSF in CD320 KO cells, suggesting that CD320 expression is necessary for the inhibitory effect (N=3, paired one-way ANOVA with Tukey’s multiplicity correction, mean +/- standard error). Panel H shows staining of human cerebellum and choroid plexus tissue with patient immunoglobulin. Immunoreactivity is seen in CD31+ endothelial cells in the cerebellum and broadly in the choroid plexus.

**Supplementary Figure 4.**
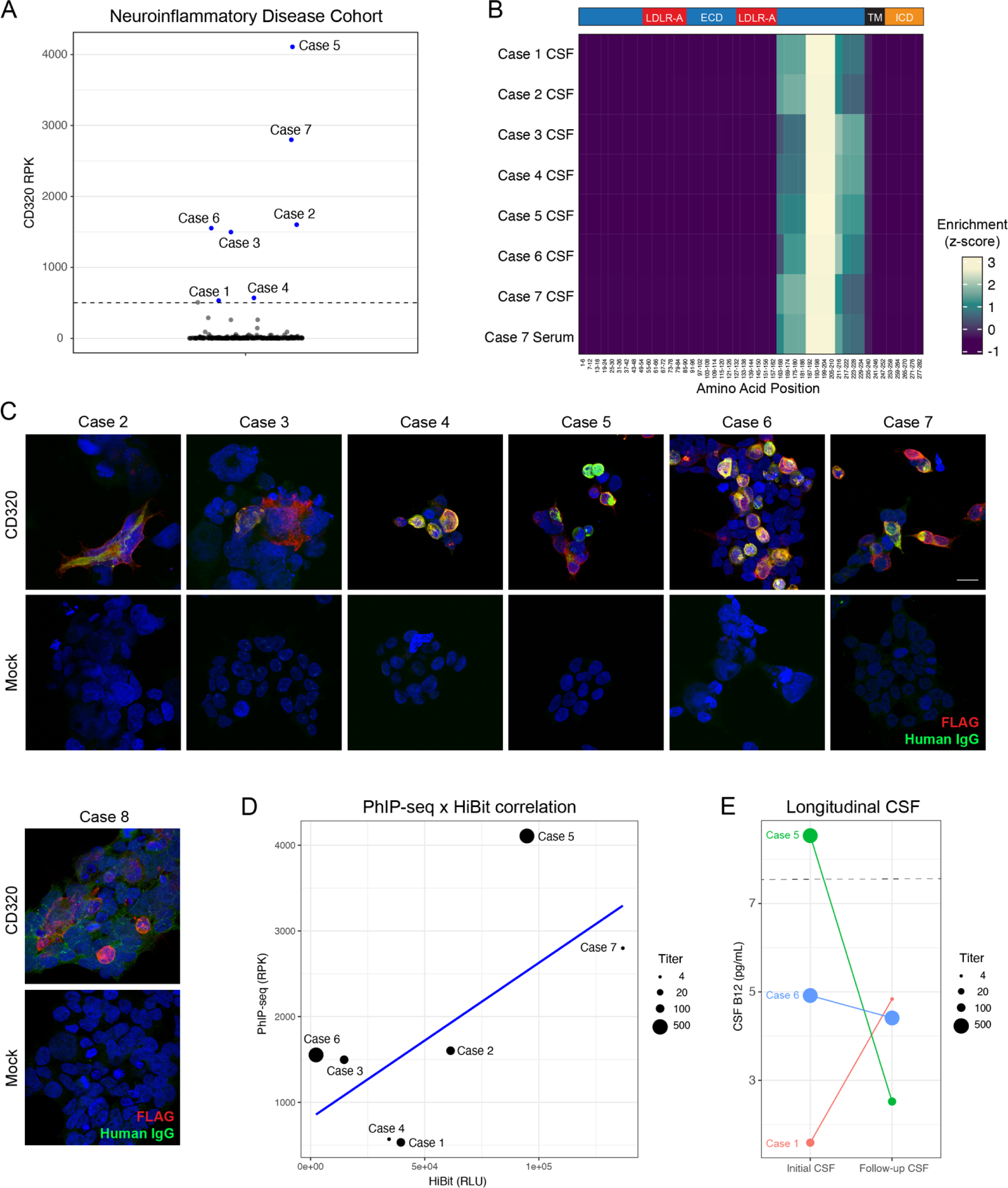
Characterization of additional anti-CD320 seropositive patients. Panel A shows CD320 enrichment from PhIP-seq experiments on 254 CSF samples from a neuroinflammatory disease cohort. Labeled samples exhibit CD320 enrichment equal to or greater than CD320 enrichment in the index patient (dotted line). Enrichment was quantified as sequencing reads per 100,000 (RPK) to account for differences in sequencing depth between samples. Panel B shows an epitope map of CD320 peptides aligned to the full-length gene (coverage is divided into five amino acid bins). Antibodies in all samples enrich the same epitope as that enriched by CSF from the index patient. Panel C shows immunoreactivity of CSF IgG for human cells overexpressing FLAG-tagged CD320. Increased immunoreactivity against the construct was observed in all cases. Scale bar, 20 microns. Panel D shows strong correlation between PhIP-seq and HiBit measurements of autoantibody target enrichment, but weak correlation to autoantibody titer as determined by cell-based assay. Panel E shows CSF autoantibody titer and corresponding CSF B12 concentration in three patients for whom longitudinal spinal fluid samples were available. In Case 1, autoantibody titers decreased from 1:100 to 1:20 after immunosuppression, and CSF B12 concentration increased, albeit in the setting of oral vitamin B12 supplementation. In Case 6, a patient who did not receive immunosuppressive treatment, autoantibody titer and CSF B12 concentration did not change with time. In Case 5, a patient without central vitamin B12 deficiency at onset, autoantibody titer decreased from 1:500 to 1:100 after immunosuppressive treatment. However, the patient developed cerebral B12 deficiency concomitant with continued neurologic decline.

**Supplementary Figure 5.**
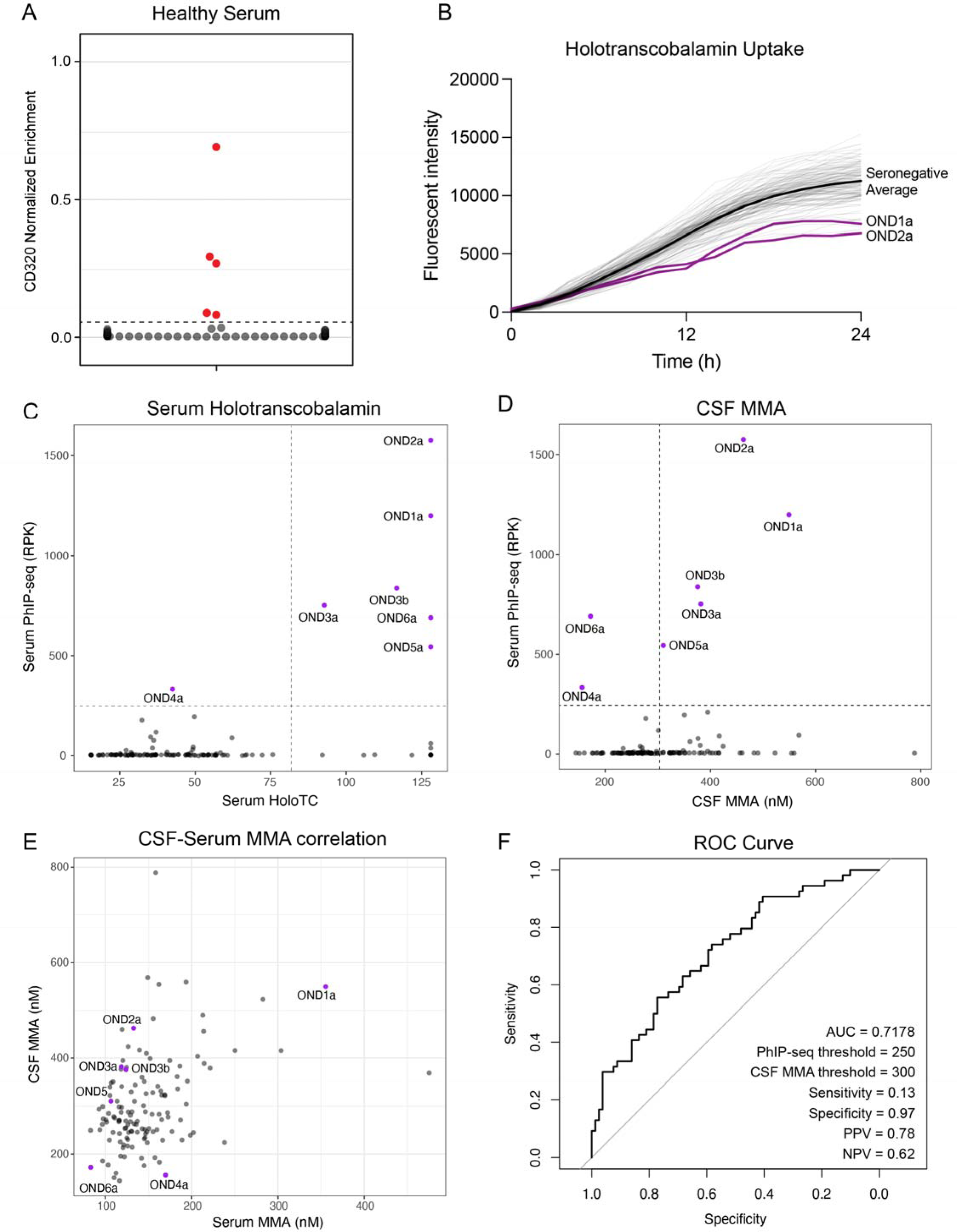
Detection of anti-CD320 in healthy controls and patients with multiple sclerosis or other neurologic diseases. Panel A shows normalized bioluminescent signal from immunoprecipitation of a tagged CD320 peptide after incubation with serum from 84 healthy controls. The dotted line corresponds to a CD320 enrichment threshold set using CSF from the index patient. Serum from five of the 84 healthy controls contained anti-CD320 autoantibodies. Panel B shows holotranscobalamin uptake by cells treated with CSF from 132 patients with multiple sclerosis (MS) or other neurologic diseases (ONDs). CSF from the two cases with the highest intrathecal anti-CD320 enrichment (purple lines) impair holotranscobalamin uptake compared to the average effect of CSF from all seronegative patients (black line). Panel C shows serum holotranscobalamin concentration vs serum anti-CD320 enrichment in patients with MS or ONDs. Holotranscobalamin accumulates in the serum of seropositive patients, presumably because its entry into the CNS is blocked. Panel D shows CSF MMA concentration vs serum anti-CD320 enrichment in patients with MS or ONDs. Panel E shows the correlation between serum and CSF MMA concentration in 132 patients with MS or ONDs. Panel F is the receiver operator characteristic (ROC) curve showing the performance of serum anti-CD320 seropositivity for predicting a high CSF MMA concentration. At an RPK threshold of 250, anti-CD320 seropositivity predicted high CSF MMA with 78% positive predictive value.

**Supplementary Figure 6.**
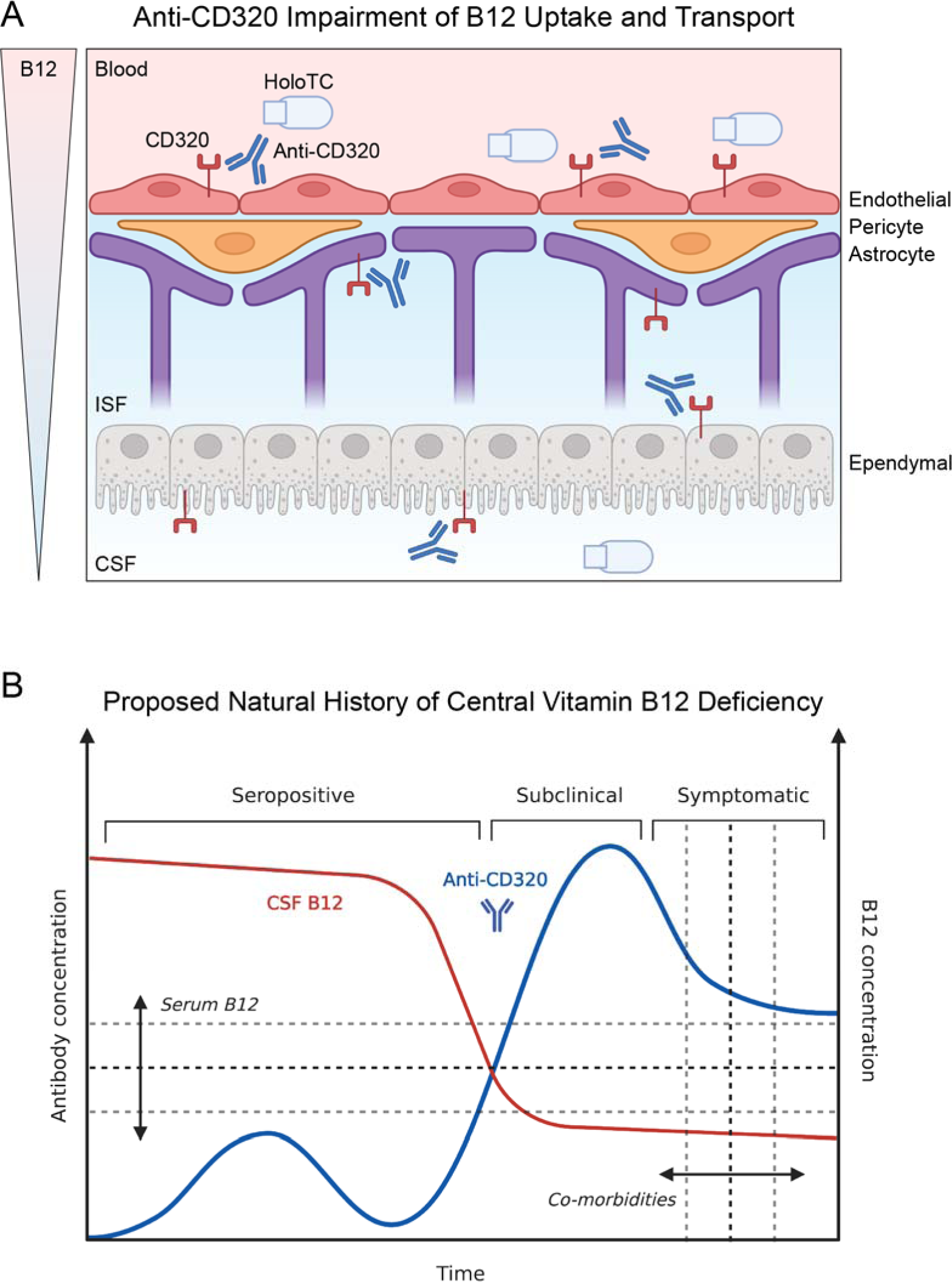
Model of anti-CD320 mechanism and natural history of disease. Panel A shows the proposed model of anti-CD320 mediated impairment of vitamin B12 transport across the blood-brain barrier. Autoantibodies in the blood impair the uptake function of CD320 on endothelial cells, possibly via internalization and depletion from the luminal cell surface. Holotranscobalamin (holoTC) must cross multiple cellular membranes on its journey to the CSF, including those of endothelial cell, pericyte, astrocyte, and ependymal cell. Anti-CD320 in the interstitial fluid (ISF) and CSF may interfere with transport at each of these borders. Furthermore, anti-CD320 in the ISF and CSF may interfere with uptake of holotranscobalamin by metabolically demanding cell types (e.g. neurons, oligodendrocytes), causing insidious neurologic sequelae. These effects likely depend on antibody titer, the duration of antibody seropositivity, and host vulnerability. Panel B shows the proposed natural history of central vitamin B12 deficiency, divided into three stages of disease (seropositive without clinical symptoms or metabolic signs, subclinical without clinical symptoms but with metabolic signs, and symptomatic with clinical symptoms and metabolic signs). The transition from seropositive to subclinical central B12 deficiency may depend on serum B12 concentration and antibody titer. The transition from subclinical to symptomatic may depend on the degree and duration of seropositivity (i.e. area under the curve) and underlying comorbidities predisposing to neurologic frailty.

