## Supplementary Appendix for "Transcobalamin Receptor Autoantibodies in Central Vitamin B12 Deficiency"

### Author Information

* Corresponding Author

### Supplementary Methods

#### Patient Enrollment and Data Collection

Patients were enrolled in a research study to detect novel autoantibodies in suspected neuroinflammatory disease (UCSF IRB# 13-12236). CSF samples were frozen and stored at -80C prior to aliquoting as undiluted CSF or diluted 1:1 in antibody sample buffer (final concentration: 20% glycerol, 20mM HEPES, 0.02% sodium azide in PBS). Healthy volunteers aged 18-65 were enrolled in a research study (UCSF IRB# 22-36117) for the collection of serum samples. Individuals were excluded if they were currently pregnant, were currently receiving immunosuppressive medications, had received chemotherapy in the last 5 years, had been diagnosed with a bleeding disorder, or had received intravenous immunoglobulin in the last 30 days. Paired serum and CSF were collected from a cohort of patients with multiple sclerosis or other neurologic diseases (IRB 14-15278).

#### Programmable Phage Display

We adapted a previously published protocol for phage immunoprecipitation sequencing (PhIP-seq). A library containing 731,724 49-amino acid peptides with 25-amino acid overlaps was cloned into T7 bacteriophage. Patient CSF was incubated with 10^10^ plaque forming units of the phage library, antibodies were enriched with protein A/G magnetic beads, and antibody-bound phage was amplified in E. coli before a second round of immunoprecipitation. Enriched phage lysates were adaptor ligated and barcoded prior to pair-end sequencing on an Illumina Novaseq to a depth of 2 million reads per sample. Reads were trimmed, aligned at the amino-acid level using RAPSearch, and normalized to sequencing depth to generate reads per 100,000 bases (RPK) for each sample. Enriched peptides were identified by calculating the fold-change of normalized counts between samples immunoprecipitated with CSF or magnetic beads only. The Benjamini-Hochberg method was used to correct for multiplicity and calculate false discovery rates.

#### 293T Cell-based Overexpression Assay

HEK293T cells were transfected with a FLAG-tagged CD320 construct packaged within the pCMV6 entry plasmid (Origene, RC200073) using Lipofectamine 3000 according to the manufacturer’s protocol. After 24 hours, cells were fixed with 4% PFA for 10 minutes at room temperature and stained with patient CSF and rabbit anti-FLAG (CST, 14793, 1:800) at 4 degrees overnight. Secondary staining was conducted with AlexaFluor-conjugated antibodies at room temperature for 1 hour. Fluorescent images were acquired on a Zeiss LSM780 confocal laser scanning microscope.

#### Western Blot

A recombinant CD320 protein containing a C-terminal GST tag and the putative epitope (Abnova, Cat. #H00051293-P01) and a recombinant truncated CD320 protein containing an N-terminal His-tag (Novus Biologicals, NBP189348PEP) were loaded into the wells of a 4 – 12% Criterion XT Bis-Tris PAGE Gel (Bio-Rad, #3450117) at 1.5ug per well. After protein separation for 1 hour at 180V at room temperature, the proteins were transferred onto Immun-Blot Low Fluorescence PVDF membranes (Bio-Rad, #1620264) using ice-cold transfer buffer containing 15% methanol and 85% 1x Tris/Glycine transfer buffer (Bio- Rad, #1610734) at 100 V for 1 hour on ice. Following protein transfer, the PVDF membranes were incubated in Intercept PBS Blocking Buffer (LiCor, [927-70001](tel:927-70001)) for 30 minutes at room temperature. The membranes were then incubated in CSF and plasma diluted in Intercept PBS Blocking Buffer at dilutions of 1:10 and 1:100, respectively, at 4°C overnight. Membranes were rinsed five times in 1X TBS containing 0.1% Tween-20 (TBS-T) and stained with goat-anti-human IRDye 800CW(LiCor, [926-32232](tel:926-32232)) at 1:10000 at room temperature for 1 hour. After five more rinses using 1X TBS-T, the membranes were imaged on a LI-COR Odyssey.

#### CRISPR-Cas9 Knockout

Primary human brain endothelial cells (Cell Systems) were cultured in Complete Classic medium in T75 flasks coated with Attachment Factor to 50% confluency. Ribonucleoprotein complexes consisting of purified Cas9 and a pool of three chemically modified single guide RNAs targeting CD320 (GGCGUCACUCACCUGCGGCC, CGGCCUCCAGGCCUAGUCCG, GGAUGGCGCAGGUUGGAGCG) were transfected into cells using the Lipofectamine CRISPRMAX Transfection Reagent according to the manufacturer’s protocol (Synthego). After 72 hours, single transfected cells were clone sorted into a 96 well plate using a BD FACSAria III. Successful knockout was confirmed by flow cytometry and immunofluorescence using a commercial anti-CD320 antibody (R&D, AF1557, 5ug/mL).

#### Holotranscobalamin Uptake Assay

Recombinant human transcobalamin II was conjugated to a pH-sensitive fluorescent dye (pHrodo Red iFL NHS ester) at a dye:protein molar ratio of 4:1. Free dye was removed using a 40 kDa MWCO Zeba Spin Desalting Column. pHrodo-conjugated transcobalamin was incubated with a 3X molar ratio of cyanocobalamin at room temperature for 1 hour to form pHrodo-holotranscobalamin. Wild type and CD320 KO HEK293T cells were grown in DMEM containing 10% FBS to 80% confluency. Cells were washed with PBS and treated with 10% healthy control CSF, patient CSF, or anti-CD320 (1ug/mL) in serum-free DMEM for 30 minutes at 37 C. 100ng of pHrodo-holotranscobalamin was added to each well, and cellular uptake was monitored every hour for 24 hours using a live-cell time-lapse fluorescent microscope (Incucyte SX5). Average total integrated orange intensity across technical duplicates was used to compare uptake dynamics between conditions.

#### Vitamin B12 ELISA

Vitamin B12 concentration in serum and CSF were measured by commercial competitive ELISA assay (Novus Biologicals) according to the manufacturer’s protocol with 2 adaptations. First, pre-coated wells were blocked overnight with sample buffer at 4 C. Second, CSF samples were diluted 1:10 prior to addition to blocked wells, and serum samples were diluted 1:100. Prior studies have demonstrated the stability of vitamin B12 with repeated freeze-thaw cycles.^[1]^

#### Holotranscobalamin ELISA

Holotranscobalamin concentration in serum and CSF was measured by a commercial ELISA assay (Tecan) according to the manufacturer’s protocol.

#### Gas Chromatography Mass Spectrometry

MMA was measured by targeted GC-MS/MS at Bevital AS (Norway). Homocysteine measurements were below the limit of detection.

#### Holotranscobalamin Binding Assay

HEK293T cells were incubated with goat polyclonal anti-CD320 (R&D, AF1557) at varying concentrations on ice for 30 minutes. Recombinant human transcobalamin was conjugated to a fluorescent dye (AlexaFluor 647 NHS ester) at a dye:protein molar ratio of 10:1. Free dye was removed using a 40 kDa MWCO Zeba Spin Desalting Column. AlexaFluor647-conjugated transcobalamin was incubated with a 3X molar ratio of cyanocobalamin at room temperature for 1 hour to form AF647-holotranscobalamin. 100ng of AF647-holotranscobalamin was added to pre-treated cells and incubated on ice for 30 minutes. Cells were washed with PBS and analyzed for fluorescent signal on a BD LSRFortessa flow cytometer.

#### Antibody Internalization Assay

Goat polyclonal anti-CD320 (R&D, AF1557) and an isotype control antibody were conjugated to a pH-sensitive fluorescent dye (pHrodo Red iFL NHS ester) at a dye:protein molar ratio of 10:1. Free dye was removed using a 40 kDa MWCO Zeba Spin Desalting Column. Wild type and CD320 KO HEK293T cells were grown in DMEM containing 10% FBS to 80% confluency. Cells were washed with PBS and treated with 1 microgram/mL of pHrodo-conjugated anti-CD320 or isotype control antibody, and cellular uptake was monitored every hour for 24 hours using a live-cell time-lapse fluorescent microscope (Incucyte SX5). Average total integrated orange intensity across technical duplicates was used to compare uptake dynamics between conditions.

#### Split-Luciferase Binding Assay

Anti-CD320 autoantibodies were detected in healthy control sera using a HiBit bioluminescent protein detection platform. A construct containing the nucleotide sequence corresponding to the previously identified CD320 autoantibody epitope and an 11-amino acid HiBit tag was used to in vitro transcribe a labeled peptide. The labeled peptide was subsequently column purified, immunoprecipitated with patient serum on Sephadex protein A/G beads, and co-incubated with a complementary LgBit peptide in the presence of luciferase substrate. Bioluminescence was measured using a Promega GloMAX plate reader.

### Supplementary Results

#### Case Vignettes

Please contact the corresponding author for access to detailed case descriptions. These descriptions have been removed to be compliant with Medrxiv privacy policies.

### Table S1. Laboratory Test Results for Case 1

| **Test** | **2014** | **2018** | **Reference** |
| --- | --- | --- | --- |
| Vit D | 31.4 |  | 10-75 pg/mL |
| Alpha Tocopherol | 13.1 |  | 4.6-17.8 mg/L |
| Ferritin | 328 |  | 11-307 ng/mL |
| Vitamin B12 | 614 |  | 180-914 pg/mL |
| MMA | 233 |  | 0-378 nmol/L |
| Hgb | 11.8 |  | 12-15 g/dL |
| MCV | 85.5 |  | 78-100 fL |
| Plt | 96 |  | 150-400 k/uL |
| TSH | 0.75 |  | 0.34-3.5 ulU/mL |
| ANA (mixed) | 1:1280 |  | <1:80 |
| RPR | Non-reative |  |  |
| Copper | 95 |  | 72-166 ug/dL |
| CSF glucose | 134 |  | 40-70 mg/dL |
| IgG index | 0.6 |  | 0-0.7 |
| OCBs | 0 |  | 0-1 |
| CSF protein | 54 |  | 15-45 mg/dL |
| CSF RBC | 2 |  | 0 |
| CSF WBC | 2 |  | 0-5 |
| CSF VDRL | Non-reative |  |  |
| Lyme IgG | negative |  |  |
| Lyme IgM | negative |  |  |
| HIV 1/2 abs | negative |  |  |
| HCV Ab | negative |  |  |
| HBV Ag | negative |  |  |
| HBV surface Ab | positive |  |  |
| Quant gold | negative |  |  |
| dsDNA antibody | 129 |  | <30 IU/mL |
| C3 | 96 |  | 71-159 mg/dL |
| C4 | 12 |  | 13-30 mg/dL |
| GAD-65 Ab | negative |  |  |
| MOG IgG |  | negative |  |
| SSA/SSB | negative |  |  |
| SM antibody | negative |  |  |
| RNP antibody | negative |  |  |
| ANCA | negative |  |  |
| Paraneoplastic Panel | |  |  |
| ANNA-1 | negative |  |  |
| ANNA-2 | negative |  |  |
| ANNA-3 | negative |  |  |
| AGNA-1 | negative |  |  |
| PCA-1 | negative |  |  |
| PCA-2 | negative |  |  |
| PCA-Tr | negative |  |  |
| CRMP-5 | negative |  |  |
| Striated muscle | negative |  |  |
| P/Q-type Ca channel | negative |  |  |
| N-type Ca channel | negative |  |  |
| AChR | negative |  |  |
| VGK channel | negative |  |  |
| NMO/AQP4 | negative |  |  |
| IgG4 | 29 |  | 1-123 mg/dL |
| TPO ab | negative |  |  |
| TG ab | negative |  |  |
| Anti-Cardiolipin IgG | 81 |  | 0-14 U/mL |
| Anti-Cardiolipin IgM | 80 |  | 0-12 U/mL |
| B2-glycoprotein IgG | 36 |  | <21 |
| B2-glycoprotein IgM | >130 |  | <21 |
| RVVT | 103 |  | 0-55.1 sec |
| HIT antibody | positive |  |  |
| ESR | 66 |  | 0-33mm/hr |
| CRP | 37.4 |  | <6.3 mg/L |
| FTA-Ab | negative |  |  |
| Cocci Ab | negative |  |  |

### Table S2. Gene and Peptide-level Counts for Phage Display

| **Gene** | **Patient 1_REP1** | **Patient 1_REP2** | **CTRL_ REP1** | **CTRL_ REP2** | **CTRL_ REP3** | **meanFC** | **FDR** |
| --- | --- | --- | --- | --- | --- | --- | --- |
| SUN1 | 6385.65025 | 2802.179481 | 3.1442247 | 1.4985302 | 6.3552737 | 1102.7134 | 0.0010749 |
| CD320 | 623.7213461 | 440.9736431 | 0 | 0 | 1.3175567 | 566.81821 | 0.0011220 |
| ALX4 | 345.0617519 | 293.3509147 | 0.1654855 | 0.0788700 | 1.9375834 | 260.08551 | 0.0014768 |
| PLAGL2 | 204.1543657 | 228.633353 | 0 | 0 | 0 | 432.78771 | 0.0014768 |
| PRRX2 | 102.9018317 | 130.8749754 | 0.0827427 | 0 | 0.3100133 | 185.26698 | 0.0095159 |
| LRRFIP1 | 3653.337712 | 2434.714076 | 15.224667 | 60.020080 | 53.244793 | 70.252403 | 0.0098385 |
| ZBTB21 | 1225.14132 | 171.5697419 | 1.7375979 | 11.830502 | 2.4026034 | 119.91884 | 0.0098385 |
| SMTNL1 | 336.8869722 | 11.06412646 | 0 | 0 | 0 | 347.95109 | 0.0174349 |
| SUGT1 | 47.68621467 | 67.97617424 | 0 | 0 | 0 | 115.66238 | 0.0252697 |
| GRM2 | 644.1582953 | 707.1189318 | 31.773218 | 8.1236113 | 6.0452604 | 42.723999 | 0.0288232 |
| MPZL3 | 25.81509366 | 129.3593416 | 0 | 0.1577400 | 0.7750333 | 95.677489 | 0.0288232 |
| ANKIB1 | 66.90411772 | 35.0869216 | 0 | 0.6309601 | 1.0850467 | 47.570346 | 0.0428992 |
| C12orf45 | 74.07497707 | 26.37202746 | 0 | 0 | 0 | 100.44700 | 0.0428992 |

| **Peptide** | **Gene** | **Sequence** | **FC** | **FDR** |
| --- | --- | --- | --- | --- |
| gi\|195972890\|ref\|NP_001124437.1 | SUN1 | PRMSRRSLRLATTACTLGDGEAVGADSGTSSAVSLKNRAARTTKQRRST | 6530.91142 | 0.00221368 |
| gi\|284925167\|ref\|NP_001165416.1 | SUN1 | SYSSDALDFETEHKLDPVFDSPRMSRRSLRLATTACTLGDGEAVGADSG | 796.077002 | 0.00372093 |
| gi\|767950082\|ref\|XP_011533022.1 | TDRP | ADPEDTVGGHPSWSGWEDDAKGSTKYTSLASSANSSRWSLRAAGRLSLK | 1728.48038 | 0.00526197 |
| gi\|55743092\|ref\|NP_068745.2 | ALX4 | RASSDLPSPLEKADSESNKGKKRRNRTTFTSYQLEELEKVFQKTHYPDV | 632.956385 | 0.00631116 |
| gi\|578805170\|ref\|XP_006712910.1 | LRRFIP1 | HESPSQDISDACEAESTERCEMSEHPSQTVRKALDSNSLENDDLSAPGR | 4527.18398 | 0.00631116 |
| gi\|740086846\|ref\|NP_001290203.1 | ACLY | MGAGKSPAGPGQKPDPGKLPAAGVLRILRGSSGLWKKRRARTSAETGRA | 680.606273 | 0.00631116 |
| gi\|767923065\|ref\|XP_011531938.1 | GRM2 | GCLFAPKLHIILFQPQKNVVSHRAPTSRFGSAAARASSSLGQGSGSQFV | 798.486228 | 0.00631116 |
| gi\|116812638\|ref\|NP_778250.2 | TDRP | EDDAKGSTKYTSLASSANSSRWSLRAAGRLVSIRRQSKGHLTDSPEEAE | 3896.71565 | 0.01358848 |
| gi\|118498345\|ref\|NP_008816.3 | ZFHX3 | PSPTKPKTKPTWRCEVCDYETNVARNLRIHMTSEKHMHNMMLLQQNMTQ | 370.918636 | 0.01438371 |
| gi\|259906403\|ref\|NP_001159367.1 | CD320 | DELGCGTNEILPEGDATTMGPPVTLESVTSLRNATTMGPPVTLESVPSV | 636.460379 | 0.01438371 |
| gi\|768004140\|ref\|XP_011526619.1 | LPPR3 | EGAPRPVAREKTSLGSLKRASVDVDLLAPRSPMAKENMVTFSHTLPRAS | 848.046847 | 0.01438371 |
| gi\|768020854\|ref\|XP_011527889.1 | ZBTB21 | SSQGSSSVSSDAPGNVLCALSQKSSLKDCSEKTALDDRPQVLQPHRLRS | 1380.29633 | 0.01438371 |
| gi\|767923088\|ref\|XP_011531949.1 | GRM2 | VSLSGSVVLGCLFAPKLHIILFQPQKNVVSHRAPTSRFGSAAARASSSL | 344.277837 | 0.01498149 |
| gi\|259906403\|ref\|NP_001159367.1 | CD320 | LESVTSLRNATTMGPPVTLESVPSVGNATSSSAGDQSGSPTAYGVIAAA | 417.039923 | 0.02154984 |
| gi\|530418157\|ref\|XP_005260493.1 | PLAGL2 | FSNGEKLRPHSLPQPEQRPYSCPQLHCGKAFASKYKLYRHMATHSAQKP | 432.181465 | 0.02154984 |

### Table S3. Additional Clinical Information for Cases 2-8

|  | **Case 2** | **Case 3** | **Case 4** | **Case 5** | **Case 6** | **Case 7** | **Case 8** |
| --- | --- | --- | --- | --- | --- | --- | --- |
| **Labs** |  |  |  |  |  |  |  |
| *B12* | 567 |  |  | 803 | 573 |  | 745 |
| *MMA* |  |  |  | **0.63 (H)** | 0.1 |  | 0.18 |
| *Homocysteine* | 6.5 |  |  |  | 10 |  |  |
| *ANA* |  | <1:80 |  | 1:40 (speckled) |  |  | <1:80 |
| *dsDNA* |  |  |  | negative |  |  |  |
| *SSA/SSB* |  | <0.2 U |  | negative |  |  | negative |
| *RF* |  | <0.2 U |  | negative |  |  |  |
| *APLAS* | negative |  |  |  |  |  |  |
| **Imaging** |  |  |  |  |  |  |  |
| *MRI brain* | Normal | Bilateral superior cerebellar peduncle T2-hyperintensity without enhancement; resolved after steroids. |  | Scattered foci of bilateral, T2/FLAIR hyperintensities in the subinsular white matter and hippocampi with two small foci of enhancement. No areas of reduced diffusion. | Multifocal white matter T2 hyperintensities |  | Normal |
| *MRI C/Tspine* | Normal | Normal |  | Multilevel degenerative changes of the spine without severe canal narrowing. | Normal |  | Abnormal cord signal within the dorsal and lateral columns of the cervical cord, and likely within the conus medullaris. |
| **Medications** |  |  |  |  |  |  |  |
| *Steroids* | IVSM | prednisone |  | prednisone | none |  | IVSM/prednisone |
| *B12* | none | none |  | 1000mcg PO daily | none |  | 2000mcg PO daily |
| *Other immune-suppression* | IVIG | Adalimumab/certolizumab for arthritis |  | Anakinra | prior rituximab for MALToma; prior R-CHOP for NHL |  | IVIG/PLEX/  rituximab |
